# Estimated Monkeypox Susceptible MSM Population in North Carolina

**DOI:** 10.1101/2022.07.21.22277860

**Authors:** Michael E. DeWitt, Mindy M. Sampson, Robert T. Fairman, Candice J. McNeil, Christopher Polk, Catherine L. Passaretti, John W. Sanders

## Abstract

Using NHANES survey data we estimate that there are nearly 65,100 North Carolina residents who identify as men who have sex with men (MSM). Among those men, it is estimated nearly 15,700 have had at least one new sexual partner in the last year and represent the highest risk for infection and onward transmission of monkeypox. Vaccination strategies should consider vaccinating with highest priority those who are highly sexually active men who have sex with men as these sexual networks have the capacity to drive the monkeypox epidemic. Estimates of the number of MSM by county as well as the estimates of highly sexually active MSM are provided by North Carolina county in order to inform vaccination quantities and priorities given the current limited supply of vaccines.

## Introduction

In May 2022, reports emerged of growing cases of monkeypox (MPX) in Europe and North America [1–3]. MPX, a double stranded DNA virus belonging to the *orthopoxvirus* genus, is endemic in West and Central Africa[4–7]. MPX is known to have several animal reservoirs which include monkeys and prairie dogs[8–10] however the landscape of potential animal reservoirs is incomplete[11,12]. Typical animal-to-human spillovers occurred through direct contact with blood, bodily fluids, or mucosal lesions on infected animals[10]. Human-to-human transmission is thought to occur from close contact with respiratory secretions, respiratory droplets, skin lesions, or fomites [13–15]. Development and climate change are bringing more humans into contact with other potential animal reservoirs which could include spillover events into the rodent population[16,17]. With the cessation of mass smallpox vaccinations, which has cross protection against MPX[7,18], it is theorized that the proportion of the population susceptible to MPX infections may have increased [19].

Early modeling studies concluded that epidemic growth outside of endemic areas would likely not be self-sustaining (i.e., basic reproduction number R_0_ < 1) [9,20–23]. Pooled estimates of outbreaks indicated a secondary attack rate between 0-11% of unvaccinated individuals with an average of 8%[24]. The case fatality rate is estimated to be between 3-6% in recent outbreaks [13]. The incubation period is estimated to be between 7-14 days with an infectious period beginning with the onset of rash until desquamation of the lesions roughly four weeks later[24,25].

The emergence of sustained human-to-human transmission, specifically in gay, bisexual, and other men who have sex with men (MSM) has shown the epidemic potential for MPX [3,26]. Furthermore, modeling of MSM sexual networks indicates that a high degree of sexual activity could continue to drive transmission with limited numbers of introductions[27]. Specifically, due to the heavy-tailed distribution of sexual partners where a small proportion of MSM have much larger numbers of sexual partners, the epidemic could grow despite having relatively low secondary attack rates (10%) [27].

Vaccines are available for MPX [28] and studies indicate that those vaccinated against smallpox have a highly reduced secondary attack rates[29]. The current guidance from the North Carolina Department of Health is to vaccinate people who are close contacts of confirmed cases (post exposure prophylaxis) and MSM or transgender individuals who have had multiple or anonymous sex partners in the last 14 days (pre exposure prophylaxis). Approaches such as ring vaccination wherein close contacts of an identified case are vaccinated could also be a useful approach for stopping onward transmission[30].

In this study we seek to quantify the population of MSM in North Carolina, where as of July 19, 2022, 20 cases have been identified. Additionally, we identify the estimated proportion of those with more than one sexual partners per year which may facilitate the spread of monkeypox [31,32].

## Methods

We utilized the 2015-2016 National Health and Nutrition Examination Survey (NHANES) to estimate the proportion of the male population in the United States who identify as a man having sex with men within the past year using the specified complex survey design [33]. This dataset also contains the reported number of sexual partners in the last year for those aged 18-69 using the survey item “In the past 12 months, with how many men have you had anal or oral sex?”. Using the 2020 Bridged Postcensal Population Estimates for North Carolina [34] we estimated the MSM population as well as the proportion who reported more than one new sexual partner in the last year.

All analysis was conducted in R version 4.1.3 (2022-03-10).

## Results

Using the NHANES survey data we find that 1.9% (95% confidence interval CI, 0.79% to 3.0%) of the United States male population identifies as a male who has sex with men. Utilizing the North Carolina male population, we further estimate that there are 65,100 (95% CI, 57,800 to 72,400) men in North Carolina who identify as MSM (Table 1, Figure 1). Of those men, we estimate that 15,700 had more than one new sexual partner in the last year (Table 1). Sexual partners amongst the MSM respondents shows a strong right-skew (heavy tailed) distribution with a small number of respondents indicating many sexual partners (Figure 2).

**Table 1:** Estimated MSM Population by North Carolina County

| County | Male Population<br>(18-69 Years Old) | Estimate MSM Population (95%<br>CI)<sup>1</sup> | Estimated MSM Population with ≥1<br>Sexual Partners in Last Year |
| --- | --- | --- | --- |
| Alamance | 53,534 | 1,010 (424 - 1,596) | 243 |
| Alexander | 12,879 | 243 (101 - 385) | 59 |
| Alleghany | 3,565 | 67 (27 - 107) | 16 |
| Anson | 8,389 | 158 (66 - 250) | 38 |
| Ashe | 8,723 | 165 (69 - 261) | 40 |
| Avery | 6,909 | 130 (54 - 206) | 31 |
| Beaufort | 14,237 | 268 (112 - 424) | 65 |
| Bertie | 6,575 | 124 (52 - 196) | 30 |
| Bladen | 10,013 | 189 (79 - 299) | 46 |
| Brunswick | 44,604 | 841 (351 - 1,331) | 203 |
| Buncombe | 85,752 | 1,617 (677 - 2,557) | 390 |
| Burke | 30,842 | 582 (244 - 920) | 140 |
| Cabarrus | 70,551 | 1,330 (556 - 2,104) | 321 |
| Caldwell | 27,144 | 512 (214 - 810) | 123 |
| Camden | 3,684 | 69 (29 - 109) | 17 |
| Carteret | 22,175 | 418 (174 - 662) | 101 |
| Caswell | 7,743 | 146 (62 - 230) | 35 |
| Catawba | 51,700 | 975 (409 - 1,541) | 235 |
| Chatham | 22,891 | 432 (182 - 682) | 104 |
| Cherokee | 8,837 | 167 (71 - 263) | 40 |
| Chowan | 4,172 | 79 (33 - 125) | 19 |
| Clay | 3,404 | 64 (26 - 102) | 15 |
| Cleveland | 31,077 | 586 (246 - 926) | 141 |
| Columbus | 17,952 | 339 (143 - 535) | 82 |
| Craven | 33,474 | 631 (263 - 999) | 152 |
| Cumberland | 112,449 | 2,121 (889 - 3,353) | 511 |
| Currituck | 9,767 | 184 (76 - 292) | 44 |
| Dare | 12,300 | 232 (98 - 366) | 56 |
| Davidson | 54,425 | 1,026 (430 - 1,622) | 247 |
| Davie | 13,543 | 255 (107 - 403) | 62 |
| Duplin | 18,116 | 342 (144 - 540) | 82 |
| Durham | 109,960 | 2,074 (868 - 3,280) | 500 |
| Edgecombe | 14,699 | 277 (115 - 439) | 67 |
| Forsyth | 119,366 | 2,251 (943 - 3,559) | 543 |
| Franklin | 24,117 | 455 (191 - 719) | 110 |
| Gaston | 72,909 | 1,375 (575 - 2,175) | 331 |
| Gates | 3,687 | 70 (30 - 110) | 17 |
| Graham | 2,623 | 49 (21 - 77) | 12 |
| Granville | 21,037 | 397 (167 - 627) | 96 |
| Greene | 8,223 | 155 (65 - 245) | 37 |
| Guilford | 170,754 | 3,220 (1,348 - 5,092) | 776 |
| Halifax | 15,234 | 287 (119 - 455) | 69 |
| Harnett | 44,835 | 846 (354 - 1,338) | 204 |
| Haywood | 19,536 | 368 (154 - 582) | 89 |
| Henderson | 35,659 | 672 (282 - 1,062) | 162 |
| Hertford | 7,917 | 149 (63 - 235) | 36 |
| Hoke | 18,261 | 344 (144 - 544) | 83 |
| Hyde | 1,911 | 36 (16 - 56) | 9 |
| Iredell | 61,198 | 1,154 (484 - 1,824) | 278 |
| Jackson | 15,089 | 285 (119 - 451) | 69 |
| Johnston | 69,817 | 1,317 (551 - 2,083) | 317 |
| Jones | 3,012 | 57 (23 - 91) | 14 |
| Lee | 19,780 | 373 (157 - 589) | 90 |
| Lenoir | 16,876 | 318 (132 - 504) | 77 |
| Lincoln | 29,265 | 552 (232 - 872) | 133 |
| McDowell | 15,217 | 287 (121 - 453) | 69 |
| Macon | 10,643 | 201 (85 - 317) | 48 |
| Madison | 7,238 | 136 (56 - 216) | 33 |
| Martin | 6,591 | 124 (52 - 196) | 30 |
| Mecklenburg | 375,767 | 7,086 (2,966 - 11,206) | 1,708 |
| Mitchell | 4,729 | 89 (37 - 141) | 21 |
| Montgomery | 8,512 | 161 (67 - 255) | 39 |
| Moore | 30,499 | 575 (241 - 909) | 139 |
| Nash | 29,907 | 564 (236 - 892) | 136 |
| New Hanover | 77,666 | 1,465 (613 - 2,317) | 353 |
| Northampton | 5,861 | 111 (47 - 175) | 27 |
| Onslow | 83,184 | 1,569 (657 - 2,481) | 378 |
| Orange | 49,709 | 937 (391 - 1,483) | 226 |
| Pamlico | 4,231 | 80 (34 - 126) | 19 |
| Pasquotank | 13,050 | 246 (102 - 390) | 59 |
| Pender | 21,244 | 401 (169 - 633) | 97 |
| Perquimans | 4,018 | 76 (32 - 120) | 18 |
| Person | 12,817 | 242 (102 - 382) | 58 |
| Pitt | 58,886 | 1,110 (464 - 1,756) | 268 |
| Polk | 6,210 | 117 (49 - 185) | 28 |
| Randolph | 47,097 | 888 (372 - 1,404) | 214 |
| Richmond | 13,906 | 262 (110 - 414) | 63 |
| Robeson | 40,345 | 761 (319 - 1,203) | 183 |
| Rockingham | 29,058 | 548 (230 - 866) | 132 |
| Rowan | 46,521 | 877 (367 - 1,387) | 211 |
| Rutherford | 21,046 | 397 (167 - 627) | 96 |
| Sampson | 19,924 | 376 (158 - 594) | 91 |
| Scotland | 11,225 | 212 (88 - 336) | 51 |
| Stanly | 20,784 | 392 (164 - 620) | 94 |
| Stokes | 14,959 | 282 (118 - 446) | 68 |
| Surry | 22,779 | 430 (180 - 680) | 104 |
| Swain | 4,313 | 81 (33 - 129) | 20 |
| Transylvania | 10,385 | 196 (82 - 310) | 47 |
| Tyrrell | 1,415 | 27 (11 - 43) | 6 |
| Union | 78,593 | 1,482 (620 - 2,344) | 357 |
| Vance | 13,108 | 247 (103 - 391) | 60 |
| Wake | 377,367 | 7,116 (2,978 - 11,254) | 1,715 |
| Warren | 6,352 | 120 (50 - 190) | 29 |
| Washington | 3,373 | 64 (28 - 100) | 15 |
| Watauga | 21,549 | 406 (170 - 642) | 98 |
| Wayne | 39,672 | 748 (314 - 1,182) | 180 |
| Wilkes | 21,779 | 411 (173 - 649) | 99 |
| Wilson | 24,936 | 470 (196 - 744) | 113 |
| Yadkin | 12,241 | 231 (97 - 365) | 56 |
| Yancey | 5,667 | 107 (45 - 169) | 26 |

Abbreviations: MSM = men who have sex with men
<sup>1</sup>95% Confidence Interval

**Figure 1:**
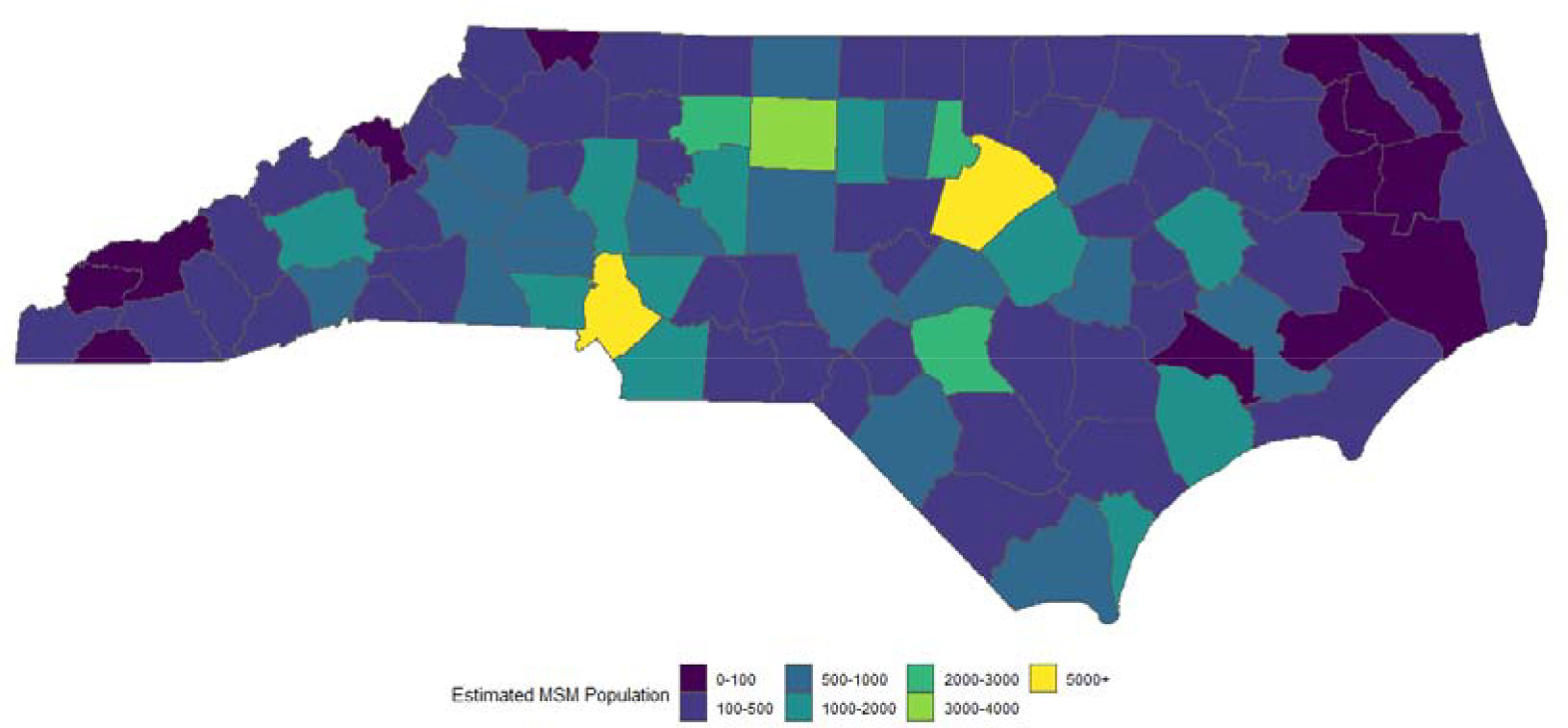
Estimated Total MSM Population by County

**Figure 2:**
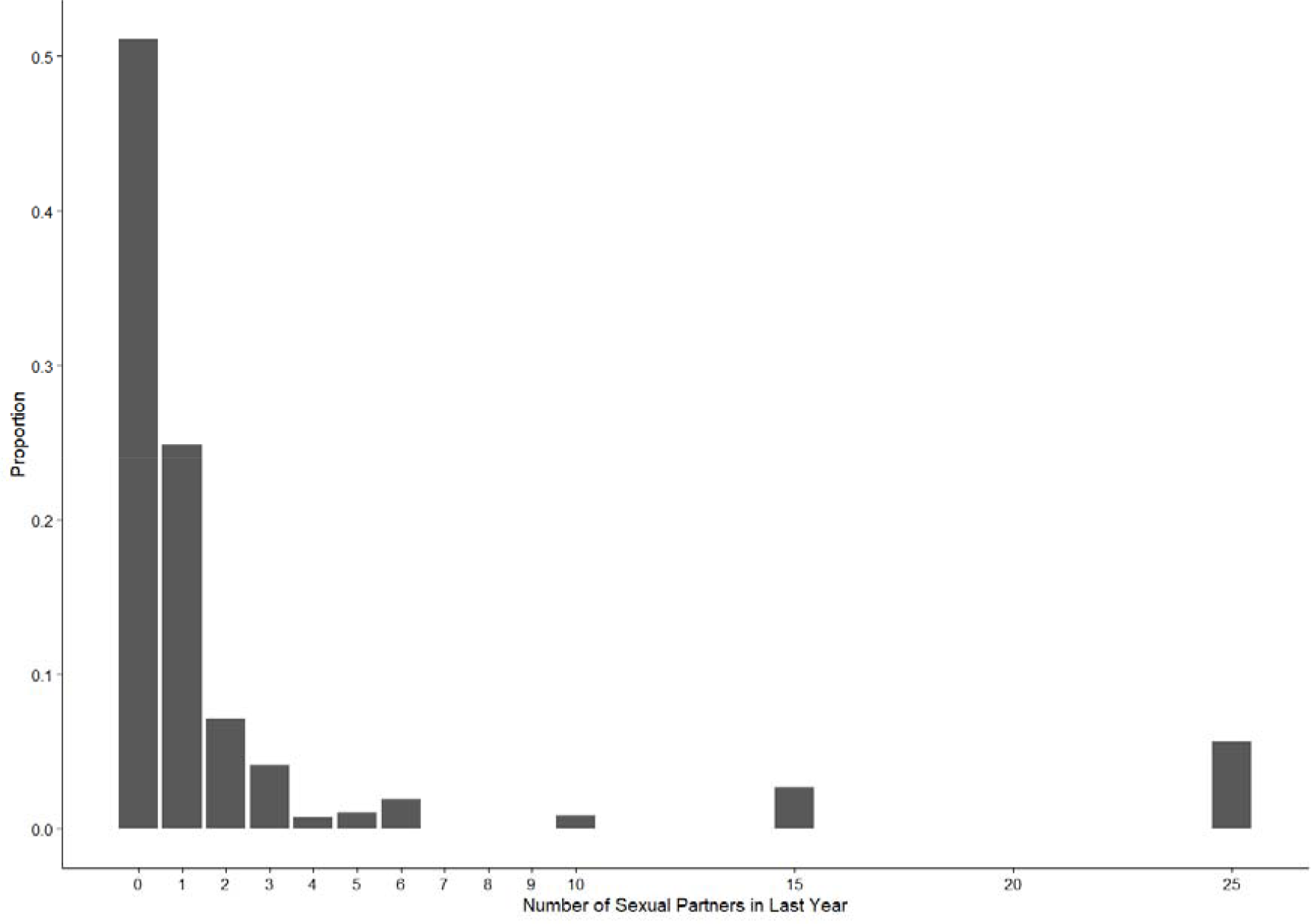
Distribution of the Reported Number of Male Sexual Partners in the Last Year Among MSM Respondents in NHANES

## Discussion

These findings suggest that at a minimum roughly 15,700 sexually active MSM in North Carolina should be offered vaccines to reduce the risk of monkeypox infection and to blunt transmission chains, which is unfortunately far greater than the current vaccine supply in North Carolina. Additionally, about 49,400 vaccines should be offered to other members of the MSM community in North Carolina to reduce their risk of infection. These estimates do not include other occupations where there is a high risk for monkeypox exposure including laboratory personnel and front-line healthcare workers with dedicated public health response roles where exposure could rapidly increase with increasing prevalence.

Outreach to those at highest risk, in this case MSM with more than one sexual partner in the last year, should be of the utmost importance. These men are at the highest risk of infection as well as could become index cases in large transmission chains. Prior work in the ongoing HIV epidemic has shown that specific outreach to those engaging in high risk activities is vital in order to slow the spread of infection[35–39].

This study is subject to several limitations. The basis for estimating sexual activity and practice relies on the accuracy of responses to the NHANES survey. Because of violence and stigma against MSM[40], respondents may not be inclined to answer questions about sexual practices, thus these estimates could underestimate the true number of MSM or the number of persons exposed to monkeypox. The NHANES survey is a national level so inferences at the subnational level may have much larger error due. These limitations are more likely to underestimate the eligible population for vaccination, suggesting significant expansion of vaccine supply will be needed in order to contain the current monkeypox outbreak.

## Data Availability

All data used to produce this analysis are available online. Additionally, the analysis will be available online at https://github.com/wf-id/mpx-at-risk.

https://wwwn.cdc.gov/Nchs/Nhanes/2015-2016/DEMO_I.XPT

https://wwwn.cdc.gov/Nchs/Nhanes/2015-2016/SXQ_I.XPT

https://www.cdc.gov/nchs/nvss/bridged_race/pcen_v2020_y20_sas7bdat.zip

## Contributions

MD, MS, RF, and CP conceived the study with input from JW. MD designed the statistical analysis plan. MD conducted the statistical analysis with input from JW. All authors had access to the underlying data. All authors contributed to interpretation of the study results. MD created the initial draft. All authors contributed to the final draft.

## Data availability

All analysis will be made available at https://github.com/wf-id/mpx-at-risk

